# Comparing the Evidential Strength for Psychotropic Drugs: A Bayesian Meta-Analysis

**DOI:** 10.1101/2021.03.04.21252887

**Authors:** Merle-Marie Pittelkow, Ymkje Anna de Vries, Rei Monden, Jojanneke A. Bastiaansen, Don van Ravenzwaaij

## Abstract

**Objective:** Approval and prescription of drugs should be informed by the strength of evidence for efficacy. While there is no formal policy towards different standards for drug approval, the typical strength of evidence might differ for different psychotropic drug groups. Using a Bayesian framework, we examine (1) whether psychotropic drugs are supported by substantial evidence (at the time of Food and Drug Administration [FDA] approval), and (2) whether there are systematic differences across drug groups.

**Methods:** Data from short-term, placebo-controlled phase II/III clinical trials for 15 antipsychotics, 16 antidepressants for depression, nine antidepressants for anxiety, and 20 drugs for ADHD were extracted from FDA reviews evaluating efficacy prior to marketing approval. Bayesian model-averaged meta-analysis was performed and strength of evidence was quantified with the Bayes factor (*BF*_*BMA*_).

**Results:** We observed substantial variation in strength of evidence and trialling between approved psychotropic drugs: Median evidential strength was extremely strong for ADHD medication (*BF*_*BMA*_ = 1820.4), but considerably lower and more frequently classified as weak or moderate for antidepressants for both depression (*BF*_*BMA*_ = 94.2) and anxiety (*BF*_*BMA*_ = 49.8). Differences might be accounted for by varying median effect sizes (schizophrenia: *ES*_*BMA*_ = 0.45, depression: *ES*_*BMA*_ = 0.30, anxiety: *ES*_*BMA*_ = 0.37, ADHD: *ES*_*BMA*_ = 0.72), sample sizes (schizophrenia: *N* = 324, depression: *N* = 218, anxiety: *N* = 254, ADHD: *N* = 189.5), and numbers of trials (schizophrenia: *Nr* = 3, depression: *Nr* = 5.5, anxiety: *Nr* = 3, ADHD: *Nr* = 2).

**Limitations:** The analysis only included pre-marketing studies.

**Conclusion:** Evidential strength varied across drug groups: Although most psychotropic drugs were supported by strong evidence at the time of approval, some drugs only had moderate or even ambiguous evidence. These results show the need for more systematic quantification and classification of statistical evidence for psychotropic drugs, and for transparent and clear communication of evidential strength toward clinical decision makers.

**Registration:** https://osf.io/5jn2d

## Background

Psychiatric disorders can be treated by various psychotropic drugs. With a wide variety of drugs available, choosing the most appropriate one can be difficult, highlighting the importance of good evidence. Clinicians must be able to trust that there is strong evidence for treatment effects. In the US, drugs must be approved by the Food and Drug Administration (FDA) before they can be marketed. Although many aspects of a drug’s profile must be considered in the approval process, the statistical evaluation of efficacy plays a central role. The FDA states that substantial evidence for efficacy is provided by “at least two adequate and well-controlled studies, each convincing on their own”.[1, p.3] Occasionally, efficacy can also be established based on “data from one adequate, well-controlled clinical investigation”[1, p.3] or existing efficacy studies of closely-related drugs, for example for modified-release (MR) variants of previously approved drugs.[1–3]

In some cases, the current statistical evaluation process may lead to suboptimal decisions. The assumption that at least two independent RCTs, or even fewer, provide substantial evidence of drug efficacy has been questioned.[4] The FDA decision process does not systematically combine the information from positive (i.e., *p*<.05, rejecting the null hypothesis of no treatment effect) and negative (i.e., *p*>.05, not rejecting the null hypothesis) trials. As such, crucial information such as the number of trials conducted before obtaining two positive trials is ignored.[5,6] Instead, the Bayes Factor (*BF*) has been suggested as a measure to quantify evidence holistically.[4,7,8] In contrast to *p*-values, *BF*s quantify evidence in favour of both the null hypothesis (i.e., no treatment effect) and the alternative hypothesis (i.e., a treatment effect) by comparing the relative likelihood of the observed data under either hypothesis.[6,9–11] For instance, a *BF*_10_ (where the subscript indicates that the *BF* quantifies the likelihood of the alternative hypothesis relative to the null hypothesis) of 30 indicates the observed data is thirty times more likely to have occurred under the alternative hypothesis than under the null hypothesis (this is considered strong evidence for the alternative hypothesis).[10] Alternatively, a *BF*_10_ of 0.2 (or 1/5) indicates the observed data is five times more likely to have occurred under the null hypothesis than under the alternative hypothesis. Finally, a *BF* around 1 indicates equipoise (i.e., the data is about equally likely to have occurred under either hypothesis).

*BFs* may not only aid drug approval but also drug prescription by clinicians. Besides effect sizes, which indicate the *magnitude* of the effect [12], strength of evidence as quantified through *BF*s indicates how likely an effect (of any positive size) is to exist. Ideally, effect sizes should be clinically meaningful and the strength of evidence sufficiently large that the drug can be considered effective with relative certainty. Previous research has shown that there are clear differences among psychotropic drug groups in terms of effect size.[13,14] There are also indications that trial programs differ between drug groups: for instance, drug approvals of antidepressants for anxiety disorders were generally supported by fewer trials than approvals of antidepressants for depression.[15,16] While, to the best of our knowledge, there is no formal policy towards different standards for drug approval, these differences may lead to differences in the typical strength of evidence for different drug groups. However, little is known about the extent to which such factors influence the typical strength of evidence for different drug groups.

### The Present Study

The goal of this study is to examine whether there are systematic differences in the strength of evidence for efficacy at approval between different groups of psychotropic drugs. We consider four major classes: antidepressants approved for depression, antidepressants approved for anxiety disorders, antipsychotics for schizophrenia, and ADHD medication. We examine whether the current evaluation process generally leads to psychotropic drugs supported by substantial evidence (at the time of approval). To determine whether there are systematic differences across drug groups in terms of strength of evidence, we compare evidential strength across the disorder groups and investigate whether trial program characteristics (e.g., effect sizes and sample sizes) are related to the strength of evidence per drug within each drug group.

## Method

This study involved publicly available trial-level data. No ethical approval was needed.

### Protocol and Registration

Study information, details regarding prior knowledge of the data, and the analysis plan were preregistered at OSFbefore data analysis but after knowledge of the data. Deviations from the preregistration are reported in the supplement.

### Data from FDA Reviews

Data on antidepressants for depression was obtained from Turner et al. [16] and de Vries et al. [17], on antidepressants for anxiety disorders from de Vries et al. [18] and Roest et al.[15], and on antipsychotics for schizophrenia from Turner et al. [14]. We extracted additional data on medications for ADHD from FDA reviews following data extraction procedures originally proposed by Turner et al. [16] described in detail elsewhere [14] and in the supplement. We also extracted additional data on antipsychotics for schizophrenia approved after publication of Turner et al. [14]. No new antidepressants were approved for depression or anxiety disorders after previous publications. In total we included data for 15 antipsychotics, 16 antidepressants approved for depression, nine antidepressants approved for anxiety, and 20 drugs approved for ADHD.^1^ For anxiety, we focused on generalized anxiety disorder (GAD), obsessive compulsive disorder (OCD), panic disorder (PD), post-traumatic stress disorder (PTSD), and social anxiety disorder (SAD). Some drugs were approved for multiple anxiety disorders. Consequently, we included 21 endorsement decisions for anxiety, resulting in a total of 72 drug-disorder combinations.

We included data for all available short-term, placebo-controlled, parallel-group and cross-over phase II/III clinical trials. We excluded studies concerned with relapse or discontinuation of the medication, long-term extension trials, and studies without a placebo control group, as these do not qualify as “well-controlled” trials [19]. We excluded data on non-approved sub-therapeutic dosages (i.e., not effective dosages), as we were concerned with the evidence load regarding dosages associated with a therapeutic effect.

### Statistical Analysis

Analysis was conducted in R(4.0.1), using the “BayesFactor” (0.9.12-4.2) [20] and “metaBMA” [21] packages.

#### Test Statistic and Effect Size Calculation

We calculated *t*-statistics using sample size and *p*-values. For parallel-group trials, we used independent samples *t*-tests and for cross-over trials, we used paired samples *t*-tests.[22] We used two-sided tests, in concordance with FDA policy. Following Monden et al. [4], we calculated a *t*-statistic for all dose levels in fixed-dose trials with multiple drug arms, while one *t*-value was calculated for flexible-dose trials with a single drug arm. When precise p-values were unavailable, t-statistics were calculated based on other information (e.g., mean differences) or imputed (see supplement). We used the standardized mean difference (SMD) as a measure of effect size. For parallel group trials, we calculated the corrected Hedges *g*. For cross-over trials, the uncorrected SMD was used.

#### Individual Bayes Factors

For each comparison, Jeffreys-Zellner Siow Bayes Factors [11,23] were calculated using *t*-statistics and sample size of the drug and placebo groups. We used a default Cauchy prior with location parameter zero and scale parameter 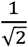.[24,25] As the FDA follows two-sided tests with a check for direction, thus de facto performing a one-sided test, we truncated below zero, and calculated one-sided *BF*s.[26]

#### Model-Averaged Bayesian Meta-Analysis

We implemented Bayesian model-averaging (BMA) [27], as neither a fixed-effect model (assuming the same underlying “true” effect-size) nor a random-effect model (being overly complex for meta-analysis of only a handful of trials) was believed to be necessarily best-suited for the present data. Instead, we weighted the results from both models according to their posterior probability, thus fully acknowledging the uncertainty with respect to the choice between a fixed or random-effect model.[28,29]

To conduct a Bayesian meta-analysis, prior distributions were assigned to the model parameters.[28] For the standardized effect size, we used a default, zero-centered Cauchy distribution with scale parameter equal to 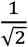.[20] For the one-sided hypothesis test, we used the same distribution with values below zero truncated. For the between-study heterogeneity parameter *τ* in random-effect models, we used an informed prior distribution based on an analysis of 14886 meta-analysis from the Cochrane Database of Systematic Reviews[30,31], namely a log normal distribution with mean *µ* = *−*2.12 and standard deviation *SD* = 1.532. We performed one Bayesian meta-analysis per endorsement decision. This yielded pooled estimates of both effect size and strength of evidence for the effects (i.e., efficacy of a certain drug for a specific mental disorder), in form of model-averaged *BF*s (*BF*_*BMA*_). For nine drug-disorder combinations supported by a single two-arm trial, we did not conduct a Bayesian meta-analysis, but used the individual *BF* and effect size instead.^2^ A total of 63 Bayesian meta-analyses was performed and results are reported for 72 drug-disorder combinations.

The resulting *BF*_BMA_ were used to describe the proportion of well-supported endorsement decisions. We used different thresholds to quantify “substantial”. A *BF*_10_ between 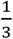 and 3 is interpreted as ambiguous evidence, while a *BF*_10_ between 3 and 10 provides moderate, a *BF*_10_ between 10 and 30 strong, and a *BF*_10_ above 30 very strong evidence for the treatment effect.[10] Importantly, these thresholds are used for demonstrative purposes and we did not aim for just another hard threshold such as *p*<.05 (see [32]).

#### Sensitivity Analysis

To study the impact of the choices we made for the prior distributions on outcomes, we performed a sensitivity analysis by setting parameter estimates varied from 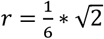 to 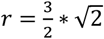.Additionally, we inspected the differences between fixed-effect and random-effect models and how excluding imputed values or cross-over trials impacted the results. Details can be found in the supplement.

## Results

### Proportion of Studies Supported by Substantial Evidence

Figure 1 and 2 visualize the results of the BMA meta-analyses for each of the disorder groups. Tables including detailed information for all analyses performed can be found in the supplement. Overall, three (4.2%) *BF*_BMA_s indicated ambiguous evidence (1< *BF*_BMA_ < 3): sertraline approved for PTSD (*BF*_BMA_ = 0.7), vilazodone (*BF*_BMA_ = 0.5), and buproprion approved for depression (*BF*_BMA_ = 2.7). Four (5.6%) meta-analytic *BF*_BMA_s indicated only modest evidence for a treatment effect (3< *BF*_BMA_ <10): Daytrana for ADHD (*BF*_*BMA*_ = 8.3), sertaline approved for SAD (*BF*_*BMA*_ = 7.3), and paroxetine (*BF*_BMA_ = 4.4) and paroxetine CR (*BF*_BMA_ = 4.4) for PD. Ten (13.9%) *BF*_BMA_s showed moderately strong evidence for treatment effects (10 < *BF*_*BMA*_ < 30): including five antidepressants approved for anxiety, two antidepressants for depression, two antipsychotics, and one ADHD medication. The majority of drugs (76.3%) indicated strong pro alternative evidence (*BF*_BMA_ > 30).

**Figure 1.**
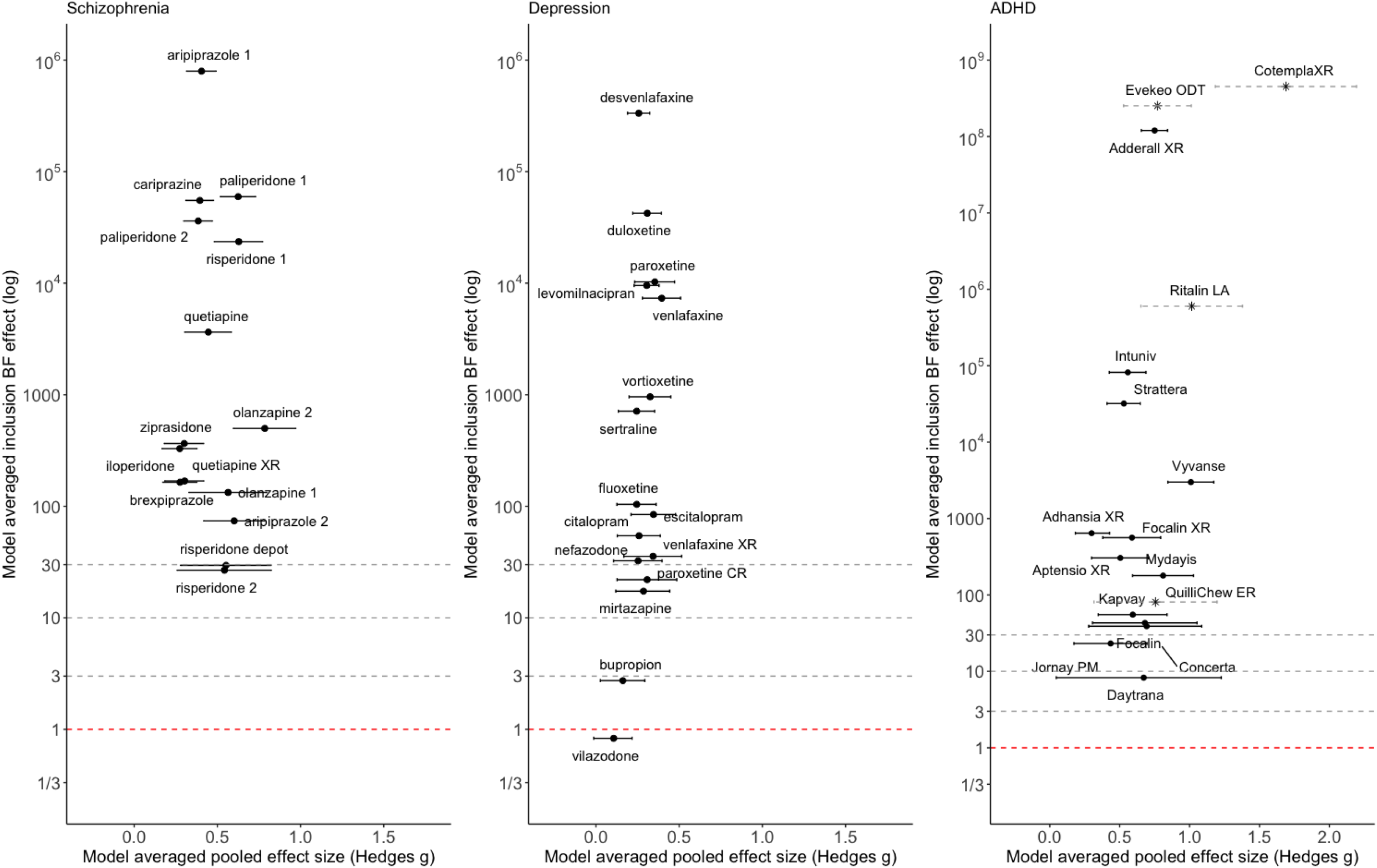
Model-averaged meta-analytic BFs and pooled effect estimates. Error bars represent 95% highest density interval. Note that the mode-averaged BF pooled effect size have different dimensions for medication approved for ADHD. For some drugs BFs and effect size correspond to a single arm trial (indicated by an asterisk, accompanied by dashed error bars indicating 95% confidence intervals). Numbers are used to differentiate drugs with the same non-proprietary name (1 = Abilify, 2 = Aristada, 3 = Zyprexa, 4 = Zyprexa Relprevv, 5 = Invega, 6 = Invega Sustenna, 7 = Risperdal, 8 = Perseris kit)

**Figure 2.**
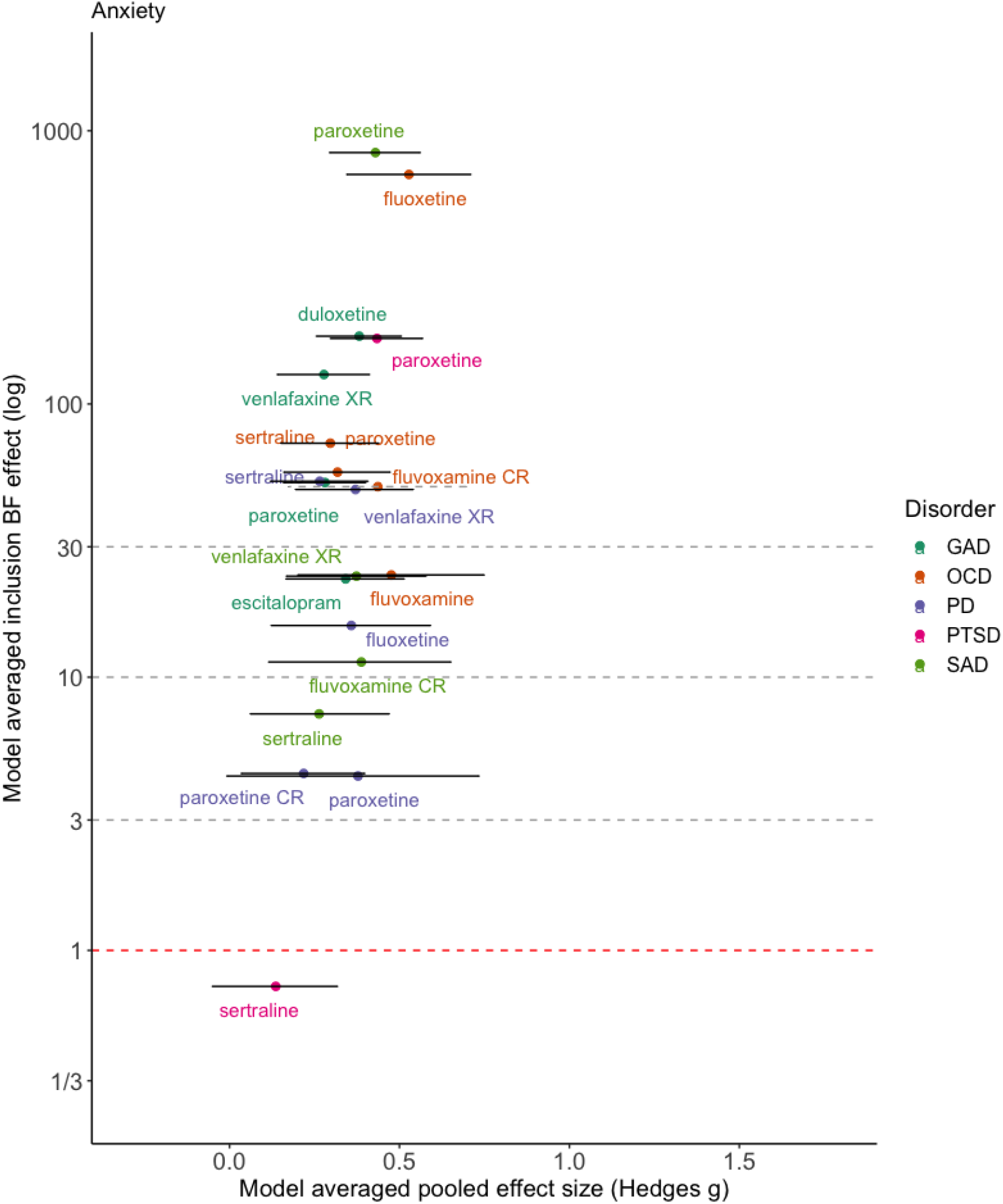
Model-averaged meta-analytic BFs and pooled effect estimates for drugs approved for anxiety disorders. Error bars represent 95% highest density interval. Colors correspond to the different anxiety disorder. For some drugs BFs and effect size correspond to a single arm trial (indicated by an asterisk, accompanied by dashed error bars indicating 95% confidence intervals).

### Differences in Strength of Evidence Across Disorders

Detailed results (i.e., meta-analytic *BF*s and pooled effect sizes, individual trial *BF*s and effect sizes, sample sizes, and number of trials) are presented in the supplement Table S1. Because the distributions of *BFs* were heavily right-skewed, we report the median instead of the mean.

The highest median strength of evidence was found for ADHD (*BF*_*BMA*_ = 1820.4), followed by antipsychotics for schizophrenia (*BF*_*BMA*_ = 365.4). Median strength of evidence was substantially lower for antidepressants for depression (*BF*_*BMA*_ = 94.2) and for anxiety (*BF*_*BMA*_ = 49.8). Although the median meta-analytic BF of each disorder group indicated “very strong evidence”, the strength of evidence differed substantially between disorders. Likewise, variability in BF_BMA_ differed between disorders. The largest variance was found in ADHD (8.3 to 2.3*10^15^), while the variance was comparable for schizophrenia (26.7 to 8.0*10^5^) and depression (0.8 to 3.3*10^5^). For antidepressants approved for anxiety the range was the smallest (0.7 to 1.3*10^5^).

### Individual *BF*s and Trial Characteristics

Median individual trial *BF*s are presented in Table S1 in the supplement (for a visual representation see Figures S1 through S4 in the supplement). There were notable differences in the individual *BF*s (i.e., *BF*s corresponding to individual trials) between the four disorder groups. The median individual *BF* was highest for ADHD (*BF* = 185.0), followed by antipsychotics (*BF* = 21.0) and anxiety (*BF* = 6.8). The median individual *BF* was lowest for depression (*BF* = 1.3). All disorder groups had enormous variation in trial strength of evidence (see Table 1).

**Table 1.** Overview of meta-analytic BFs (BF_BMA_) and pooled effect sizes per drug (ES_BMA_), individual trial BFs (BF) and effect sizes (ES), sample size for individual trials (N_i_), and number of trials (N_trials_) across the four disorder groups.

|  |  | Schizophrenia | Depression | Anxiety | ADHD |
| --- | --- | --- | --- | --- | --- |
| $BF_{BMA}$ | Median | 365.4 | 94.2 | 49.8 | 1820.4 |
|  | range [min-max] | 26.7 - 8.0*10<sup>5</sup> | 0.83- 3.3*10<sup>5</sup> | 0.7 - 1.3*10<sup>5</sup> | 8.3 - 2.3*10<sup>15</sup> |
| $ES_{BMA}$ | Median | 0.45 | 0.30 | 0.37 | 0.72 |
|  | range [min-max] | 0.27 - 0.79 | 0.11 - 0.40 | 0.14 - 0.55 | 0.30 - 1.89 |
| $BF$ | Median | 21.0 | 1.3 | 6.8 | 185.0 |
|  | range [min-max] | 0.1 - 1.0*10<sup>9</sup> | 0.1- 7.5*10<sup>7</sup> | 0.1 - 4.6*10<sup>64</sup> | 0.2 - 1.4*10<sup>19</sup> |
| $ES$ | Median | 0.44 | 0.27 | 0.38 | 0.65 |
|  | range [min-max] | -0.13 - 0.92 | -0.29 - 0.84 | -0.15 - 1.15 | 0.04 - 1.89 |
| $N_i$ | Median | 324 | 218 | 254 | 189.5 |
|  | range [min-max] | 68 - 636 | 29 - 704 | 87 - 565 | 20 - 563 |
| $N_{trials}$ | Median | 3 | 5.5 | 3 | 2 |
|  | range [min-max] | 1 - 6 | 3 - 16 | 1 - 4 | 1 - 6 |

The relationships between individual *BF*s and both effect size and sample size are shown in Figure 3. The scatterplot of effect size and individual *BFs* shows a positive association, whereas the scatterplot of sample size and individual BF does not show a clear pattern (e.g., the highest *BF* for ADHD trials had the smallest sample size), although this may be due to confounding with other factors (such as effect size).

**Figure 3.**
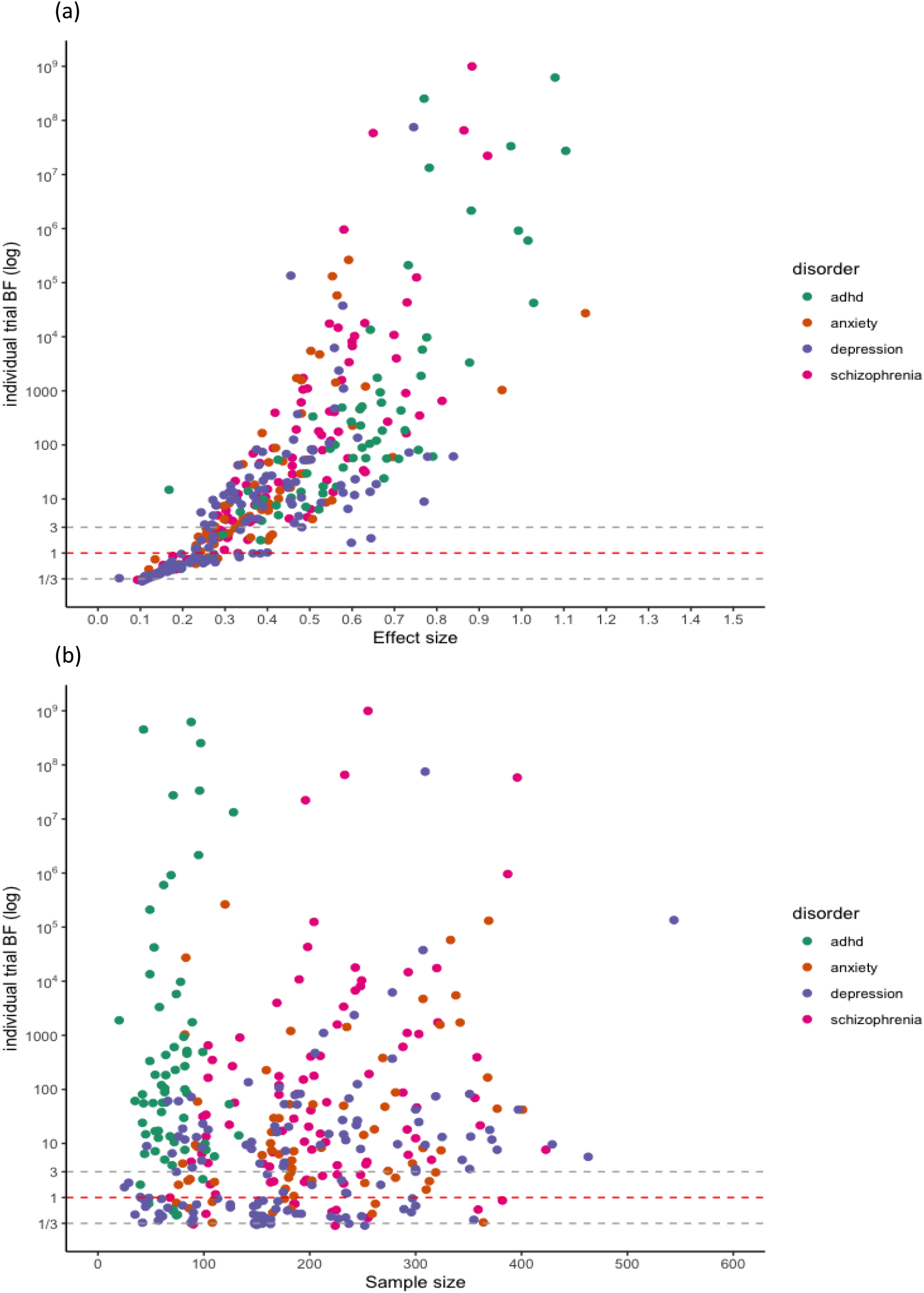
Individual BFs plotted against effect size (top) and sample size (bottom). Colours indicate the four different disorder groups.

We observed substantial variation in sample size for trials concerning ADHD medication (20 to 563). This can be partially explained by the inclusion of cross-over trials, which by design have comparatively small sample sizes. Trials were generally randomized, controlled, parallel group trials (parallel-group RCTs); however, crossover trials were sometimes performed for drugs approved for the treatment of ADHD. Notably, smaller sample sizes did not necessarily correspond to ambiguous evidence. For instance, two cross-over trials for ADHD indicated very strong pro-alternative evidence with small sample sizes (*n*=20, *ES* = 1.74, *BF* = 189.4 and *n* = 39, *BF* = 3.70*10^14^). This was the case for parallel trials for depression and schizophrenia as well (*n*=66, *ES* = 0.84, *BF*=61.1 and *n*=104, *ES* = 0.73, *BF*=163.7).

The lowest individual *BF*s were found for depression (*BF* = 1.3), which might be explained by the small effect sizes (*ES* = 0.27), as illustrated in Figure 3(a). In contrast, antipsychotics and ADHD medication showed larger effect sizes (*ES* = 0.44 and *ES* = 0.65, respectively) corresponding to stronger evidential strength.

### Meta-Analytic *BFs* and Trial Characteristics

The very strong evidence obtained for most ADHD medications appeared to be primarily due to high individual *BF*s, as the number of trials for each ADHD drug was very small. In contrast, very strong evidence for depression was generally achieved through a large number of trials, despite small studies and very low individual *BF*s. Notably, larger numbers of trials corresponded to a greater proportion of trials being deemed questionable or negative by the FDA. For example, for paroxetine for depression, 16 trials were mentioned, of which nine were deemed questionable or negative. Our Bayesian re-analysis suggests evidence for an additional trial to be ambiguous. Nonetheless, the meta-analytic *BF* suggested very strong pro-alternative evidence for the drug to treat depression (*BF*_*BMA*_ = 10267.8).

Under a Bayesian framework, more trials (i.e., more data), equal more evidence for the more probable hypothesis.In other words, with accumulating data the evidential strength (i.e., the *BF*) tends to point towards either zero (in case the null hypothesis is true) or infinity (in case the alternative hypothesis is true). However, we do not observe this relationship across drugs in practice (see Figure 4). A likely explanation is that more trials are run for drugs with lower effect sizes to compensate. Trials concerning antidepressants approved for anxiety were slightly higher powered (i.e., had larger effect sizes and sample sizes) compared to those for antidepressants approved for depression. Nonetheless, only few trials were performed per drug resulting in weaker evidence at the drug level compared to the other three disorder groups. For antipsychotics, we observed substantial pro-alternative evidence across the board while the number of trials was comparable to those for antidepressants for anxiety. However, similar to ADHD medication, the individual studies were on average well-powered (i.e., medium effect size and large sample size) resulting in higher individual *BF*s and consequently stronger evidence at the drug level.

**Figure 4.**
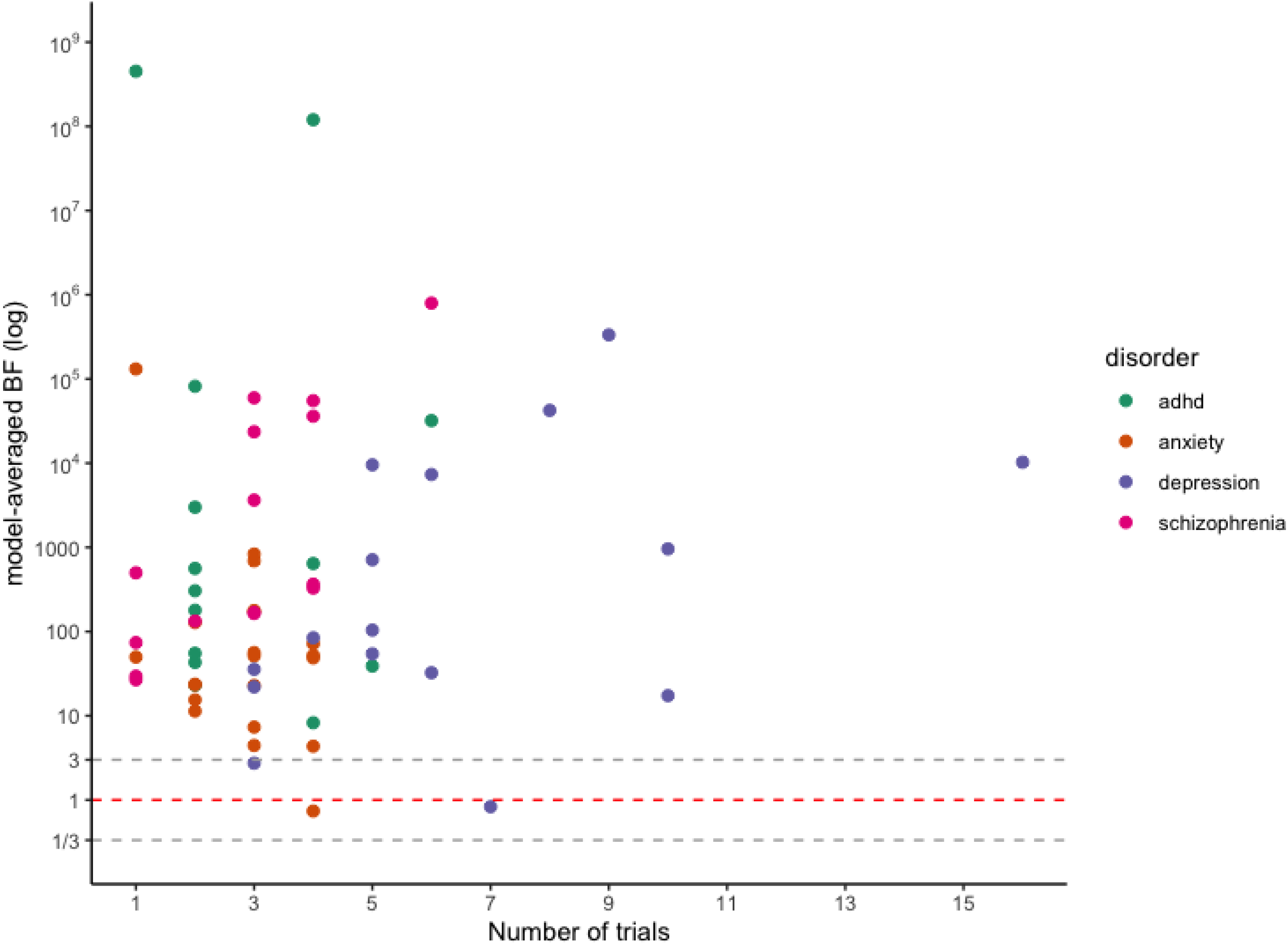
Model-averaged BFs plotted the number of performed trials. Colours indicate the four different disorder groups.

### Sensitivity Analysis

Results from the sensitivity analyses are summarized in Table 2 in the supplement. Importantly, the qualitative interpretation did not change for different choices of model and/or scale parameter.

## Discussion

Even though approval of psychotropic drugs is based on the same guideline and processed through the same pathway and by the same group within the FDA, we detected large differences in evidential strength and trial programs. While efficacy for the majority of psychotropic drugs was supported by very strong evidence at the time of approval, we observed substantial variation in the strength of evidence between approved psychotropic drugs: ADHD medication was supported by extreme evidence, whereas evidence for antidepressants for both depression and anxiety was considerably lower and more frequently classified as weak or moderate.

Differences in evidential strength might be partly explained by differences in trial programs. For instance, ADHD drugs typically had very large effect sizes, resulting in extreme evidence for efficacy despite comparatively fewer and smaller trials. All else being equal, larger effect sizes correspond to larger *t*-values, which in turn corresponds to larger *BF*s. A potential drawback here is that the drug is tested on too few people to effectively gather evidence to rely on for safety. As most ADHD drugs are variants of methylphenidate, this may be considered acceptable. However, one might wonder: if a drug is considered different enough that a new approval application with new trials is needed to establish efficacy, is it reasonable to assume that safety will be the same?

In contrast, for depression in particular we saw clinical trial programs with comparatively many trials and participants, meaning that there is much more experience with the drug at the time of approval. Evidence for efficacy, however, is considerably lower compared to ADHD and schizophrenia. The most likely explanation for this finding is that effect sizes for antidepressants were generally smaller than for other drug groups. Alternatively, heterogenous samples for depression and anxiety, due to more ambiguous diagnostic criteria, might have contributed to larger between-study variation and thus lower evidential strength.

Using a Bayesian approach allowed us to identify cases in which psychotropic drugs were approved with moderate or even ambiguous evidence for efficacy. Approximately a quarter of all meta-analytic Bayes factors fell within this tier (i.e., *BF*_BMA_ < 30). In a few instances, drugs were approved despite ambiguous statistical evidence (i.e., 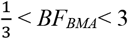). Sometimes, approval was based on other considerations. For example, bupropion SR (sustained-release) was approved based on bio-equivalence with immediate-release bupropion, despite negative efficacy trials for bupropion SR.[33] Other times, negative or “failed” trials were not included in the efficacy determination. For example, for vilazodone, three of five trials were considered “failed”, as the active comparator did not separate from placebo. The FDA has a history of ignoring failed trials because they supposedly lack assay sensitivity, the ability to differentiate an effective treatment from a less effective or ineffective one.[34] While other considerations certainly play a role in the approval process, the example of vilazodone illustrates how the FDA’s current practice of determining efficacy using two independent statistically significant trials (regardless of the number of additional negative trials) *can* lead to inconsistent decision making in practice. Under the Bayesian framework, endorsement of this drug would not have been recommended.

### Strengths and Limitations

Adopting a Bayesian framework enabled us to capture differences in evidential strength between disorders and drug groups. Nonetheless, the results should be considered in light of a few limitations. First, while we used information from FDA-registered trials, limiting the influence of reporting bias, we were confined to data from approved drugs and pre-marketing studies. Consequently, we cannot speak to the process and statistical evidence of non-approval, or strength of evidence after post-marketing studies. Second, some values were unavailable and had to be imputed, which might have introduced extra noise. Nonetheless, imputation did not seem to be associated with increased between-study heterogeneity as indicated by comparable posterior probabilities of the random-effect model between drugs for which test statistics were available and drugs for which test statistics were imputed. Lastly, Bayesian analysis is dependent on the choice of prior. While this is an often-heard critique, we mostly restricted our analyses to default priors to ensure comparability of our results across drug groups. Furthermore, our sensitivity analysis indicated that different choices for the scale parameter of the prior did not change interpretation of the *BF* qualitatively in the present analysis.

A main strength of the study is that, to our knowledge, we are the first large-scale comparison of evidential strength between several disorders. Previously, Bayesian methods have been proposed for and discussed in the context of the drug development and endorsement.[4,6,7,35–39] In recent years, Bayesian network meta-analyses specifically have become increasingly popular in medical sciences.[37,38,40,41] Our work differs in several respects. Unlike previous research, we were not concerned with personalized treatment for a single disorder. Moreover, while previous investigations were concerned with efficacy and tolerability, they either did not speak about evidential strength or did not compare evidential strength across different psychological disorders. Here, we provided an overview of the evidential standard for psychotropic drugs at the time of FDA approval and demonstrated how psychotropic drugs differ in their evidential strength, using *BFs*.

### How Bayes Factors Could Aid Evidence-Based Treatment Choices

For the purpose of drug development and endorsement, Bayesian meta-analysis offer several advantages over classical, frequentist meta-analysis, suggested by the FDA.[42] While frequentist meta-analysis is well-equipped to estimate the size of a treatment effect and its uncertainty [6], it cannot differentiate between absence of evidence (uncertainty regarding the effect) and evidence of absence (e.g., evidence for effect = 0; a similar argument is made by [4]). This is especially important in the context of failed or negative trials, which could either indicate insufficient data or non-effectiveness of the drug. If the problem is merely absence of evidence, the sponsor might perform additional trials to prove efficacy, while non-approval should be issued when evidence of absence has been demonstrated.

For the purpose of drug prescription, *BF*s offer a valuable source of information for clinicians. Prescription and use of psychotropic drugs has steadily increased over the past decades.[43–45] With a wide variety of drugs available, choosing the most appropriate one can be difficult, highlighting the importance of good evidence. Next to safety and patient-specific concerns, considerations regarding effect size and evidential strength play a central role. Commonly, strength of evidence is assessed by using qualitative or subjective criteria. For example, the APA considers evidential strength for their recommendations by reviewing the available literature and assessing risk of bias, the degree to which reported effects point in the same direction, directness of the outcome measure, quality of the control condition, and precision of the estimate (typically through width of a 95% confidence interval).[44] While these considerations are meaningful, implementation can be unsystematic, easily influenced by the rater, and might fail to effectively quantify strength of evidence (i.e., the likelihood of the treatment effect existing). For example, the APA recommends sertraline for the treatment of PTSD and argues that this decision is supported by moderate strength of evidence. In contrast, our analysis suggests no evidence for a treatment effect of sertraline at the time of approval and implies that it was more likely that sertraline does not alleviate PTSD symptoms.

Moreover, *BF*s offer a valuable source of information in cases where effect sizes are highly comparable. For instance, the APA concludes that many antidepressants are equally effective [46] and makes no clear recommendation which one to prefer. In these cases, *BF* could be used as an additional criterion, as antidepressants vary substantially in evidential strength (see also [4]). For example, leaving aside non-efficacy considerations, but considering both the effect size and evidential strength, one might choose venlafaxine or paroxetine over sertraline or citalopram, two very commonly prescribed antidepressants.[48] This advantage still holds if effect sizes vary from medium to large. For example, for ADHD drugs, Cotempla XR, Evekeo ODT, and Adderall clearly demonstrated the highest evidential strength with comparable effect size and might be preferred over the others.

## Conclusion

Taken together, the present analysis offers interesting insights into the evidential strength within and across different psychotropic drugs. We observed large differences in evidential strength and trialling between disorders. While the majority of re-analysed drugs was supported by substantial evidence, we also observed cases where the current approval process led to endorsement despite ambiguous statistical evidence. Moreover, evidential strength differed greatly between drugs and across disorder groups. Lower evidential support for efficacy was observed more frequently for antidepressants. Differences in evidential strength might be a consequence of different standards in trialling. The Bayes factor as a measure of evidential strength might offer a valuable, additional source of information and help to set up a consistent and transparent standard for evaluating strength of evidence of efficacy in the approval process of psychotropic drugs.

## Data Availability

The datasets generated and analysed during the current study are available at OSF, https://osf.io/364t5/

## Contribution

Initial planning and preregistration were drafted by DvR, YAdV, and MMP, and RM and JAB provided feedback. YAdV and MMP collected the additional data. MMP performed data analysis and drafted the manuscript under supervision of DvR and YAdV. All co-authors provided detailed feedback. The corresponding author attests that all listed authors meet authorship criteria and that no others meeting the criteria have been omitted

## Acknowledgement

We would like to thank the Center for Information Technology of the University of Groningen for their support and for providing access to the Peregrine high performance computing cluster. Rei Monden was partially supported by the Clinical Investigator’s Research Project in Osaka University Graduate School of Medicine.

## Footnotes

1 For reporting of ADHD drugs, we use the commercial instead of the non-proprietary names as they are commonly variants of the same active agent.

2 That is, fluvoxamine CR approved for OCD, paroxetine CR approved for SAD, and seven drugs approved for the treatment of ADHD (i.e., Contempla XR, Daynavel XR, Evekeo ODT, Metadate CD, QuilliChew, Quillivant, and Ritalin LA).

